# The Effect of an 8-Week Pilates Training Program on Flexibility and Sport-Specific Performance in Young Female Artistic Swimmers

**DOI:** 10.64898/2025.12.18.25342564

**Authors:** Lydia Kainourgiou, Stavroula Ntomali, Theodoros M. Bampouras, Maria Karydaki, Eleni Dimakopoulou

## Abstract

**Purpose:** Female artistic swimmers’ successful competitive performance depends on several physical performance factors such as postural control, flexibility, muscle strength, and aerobic endurance. Flexibility is a highly important physical component that can directly impact successful performance. Pilates could be an appealing training modality to artistic swimmers due to the sport’s reliance on precise body control and synchronized movements. This study aimed to assess the impact of an 8-week Pilates training program on flexibility and sport-specific performance in young female artistic swimmers.

**Methods:** Eighteen competitive artistic swimmers aged 13 to 15 years (13.8±0.8 years) were randomly assigned to an experimental or control group. The experimental group executed a Pilates class, incorporating typical equipment exercises (small Pilates ball and bands), while the control group maintained their regular gym workout routine. Both groups had two training sessions per week for 60 minutes per session. Every athlete was evaluated on their ability to accomplish two basic figures in the water (Ariana, Rio). To evaluate all characteristics, univariate analyses of covariance (ANCOVA) were employed, with initial measurements as covariates and final measures as dependent variables.

**Results:** Flexibility exercises, splits (*P* =.028), bridge (*P* =.003), shoulders (*P* =.005), and figures (Ariana: *P* =.001, Rio: *P* =.003), demonstrated statistically significant differences in favour of the experimental group. However, there was no difference in knee flexibility across the groups (p=0.376). The covariate had a significant impact (p<.05) across all analyses.

**Conclusion:** The findings suggest that Pilates training enhances flexibility and basic figure performance in young female artistic swimmers. Incorporating Pilates into training programs could be beneficial in improving flexibility and overall performance.

## Introduction

Artistic swimming (AS) is an aesthetic sport, with performance success determined by the athlete’s flawless execution of various figures while keeping in time to music, as that returns the highest technical score. The competitive structure of AS is organized by World Aquatics, the governing body for all aquatic sports, which defines the technical requirements of the sport. These requirements include compulsory figures, specifying body positions, movements, and transitions, which athletes must execute with precision. The sport continually evolves, incorporating increasingly challenging lifts, throws, and acrobatic skills (Mountjoy et al., 2010). Consequently, AS combines dance, acrobatics, and various swimming styles and demands mastery in cardiorespiratory fitness, balance, coordination, agility, swimming proficiency, explosiveness, endurance, and musical expression (Chairopoulou, 2010). Both in-water and dry (e.g. gym) programmes routinely train these physiological characteristics (Dodigović & Sindik, 2015, Tinto, Campanella, & Fasano, 2016).

Pilates is a holistic training approach that emphasizes mental and physical control, optimal breathing, flexibility, strength, and precision (Ahearn, 2006; Isacowitz, 2022). Created by Joseph Pilates and initially referred to as “Contrology,” Pilates integrates principles from gymnastics, martial arts, yoga, and dance into a series of 34 exercises, providing several benefits such as strength, muscular endurance, flexibility, changes in body composition, and balance (Latey, 2001; Siler, 2000). Pilates is an ideal training technique that reduces the risk of injury due to its low-impact nature and documented positive effects on flexibility, strength, and postural control, all of which are essential for athletic performance (Hornsby & Johnston, 2020). Indeed, the benefits of Pilates have been well documented across various sports. For example, professional dancers and soccer players have demonstrated improved strength, flexibility, and range of motion when Pilates was incorporated into their training regimens (Ahearn, 2006; Amorim, Sousa, & Santos, 2011; Chinnavan, Gopaladhas, & Kaikondan, 2015). Similarly, Pilates training has been found to enhance balance, muscle responsiveness, and performance in dynamic movements like jumps and sprints for athletes in volleyball and basketball (de Costa Ventura, 2020; El-Sayed, Mohammed & Abdullah, 2010).

With its emphasis on core strength, flexibility, and controlled movements, Pilates appears particularly suited to artistic swimmers due to the sport’s reliance on precise body control and synchronized movements. Flexibility plays a key role in ensuring fluidity in routine execution and enabling complex maneuvers like lifts, throws, and intricate transitions by allowing greater range of motion and adaptability (Boyle & Verstegen, 2012; Hornsby & Johnston, 2020) potentially increasing the score achieved. Increased flexibility has been linked to better AS performance (Yamamura et al., 1999) and has also been reported as the most important physical component for performance in AS (Cho et al., 2017) .. The enhanced flexibility and joint mobility achieved through Pilates could be essential for the successful execution of the wide range of movements required in AS routines (Badau, Szabo-Csifo, Ciulea, Alexandrescu, & Badau, 2021; Cho et al., 2017; Wells, Kolt, & Bialocerkowski, 2012).

Despite the potential benefit of Pilates in AS, the use of Pilates in this sport has yet to be examined. Given the increased technical demands (Chu, 1999) and the complexity of this sport and the search for varied training methods to improve performance (Ponciano et al., 2018), the purpose of the current study was to explore how Pilates training affects the flexibility and sport-specific performance of young female artistic swimmers.

## Methods

### Participants

The study involved 18 AS athletes from a local club, aged 13 to 15 years (13.8±0.8 years), with a height of 159.5±6.0 cm and a body mass of 52.4±9.1 kg. Participation prerequisites included a minimum of two years of experience in AS and active athletic engagement for two years. All participants and their parents/guardians received comprehensive information about the study’s objectives, procedures, and requirements. A signed consent form from the athletes’ parents/guardians was obtained, confirming voluntary participation and confidentiality, with the option to withdraw at any time. The study received approval from the University’s Ethics Committee (Protocol No: 1461/11-01-2023).

### Procedures

Before the initiation of the intervention program, athletes underwent initial assessments (PRE) of flexibility and sport-specific figures (see Measurements below). Following a 15-minute warm up of static and dynamic stretching exercises (e.g. seated forward pose, lunge stretch, pigeon pose, cat-cow stretch, camel pose, shoulder rolls etc.), flexibility was assessed in the University gym (REMOVED FOR REVIEWING PURPOSES). Figure performance was scored by independent certified judges at the 25m indoor swimming pool of (REMOVED FOR REVIEWING PURPOSES), with an average water temperature of 27°C.

Subsequently, the athletes were randomly assigned (using block randomization; sealedenvelope.com) to either the control group (CON, n = 9) or the experimental (EXP, n = 9). CON adhered to the usual gym training routine as planned by the club’s main coaches, following their annual periodized schedule. EXP replaced their usual gym training routine with a Pilates programme comprising of movements from the classic Pilates repertory, with adjustments using small props (e.g., small Pilates ball, Pilates bands). The training program duration was eight weeks, with two training sessions per week for 60 minutes per session for both groups. Both groups continued with the same and planned in-water training. All participants were required to attend at least 80% of the training sessions without missing two consecutive sessions, both in and out of the water. The researcher conducted the measurements under the supervision of the athletes’ main coaches and all evaluations (flexibility and figures) were repeated at the end of the eight-week intervention (POST).

### Measurements

#### Flexibility

All flexibility measurements were conducted on the ground using a basic 2-meter tape measure and were taken in centimeters (cm) for four tasks: a) Split (right, left, and middle, measuring pubic bone distance from the ground, with a lower score indicating higher flexibility (pubic bone closer to the ground)), b) Bridge (full bridge, measuring palm-to-heel distance; with a lower score indicating higher flexibility (palms closer to the heels)), c) Shoulder flexibility (measuring the distance between palms after a few shoulder circumferences while holding a sturdy object, such as a belt, gradually reducing the grip width until reaching the minimum possible distance that still allows full rotations; thus, a lower score indicating higher flexibility (palms closer together)), and d) Knee extension (the athlete sits in a seated position and extends the knees as much as possible while maintaining posture). The measurement is taken by calculating the gap between the lifted heels and the ground, which reflects the degree of knee extension. Α higher score reflects greater improvement in knee extension, as it corresponds to an increased ability to fully extend the knees while maintaining the required posture. All tests were conducted after a standardized warm-up, and all measurements were recorded after a single trial.

Sport-specific figures

The figures selected for evaluation were Ariana (423DD 2.2) and Rio (143DD 3.1) [39], as these figures require strength, flexibility, precision, control and fluidity of movement. Ariana (Figure 1) commences with the swimmer lying flat on their back in the water, followed by an arched back with the upper body in the water. One leg is then lifted in a 180° arc over the surface of the water and stops in a split position. With the swimmers’ legs in that position, the hips rotate 180° and the swimmer executes a walkout front, returning to the starting position facing the opposite direction. Rio (Figure 2) also commences with the swimmer lying flat on their back in the water but has a more complex skill execution. The knee, shin and toes of the first leg are drawn along the surface of the water before it is straightened and followed by the other leg. The swimmer then submerges and subsequently thrusts themselves upwards, with their feet and body in a vertical position (akin to a ground handstand), while a spin concurrently performed.

**Figure 1.**
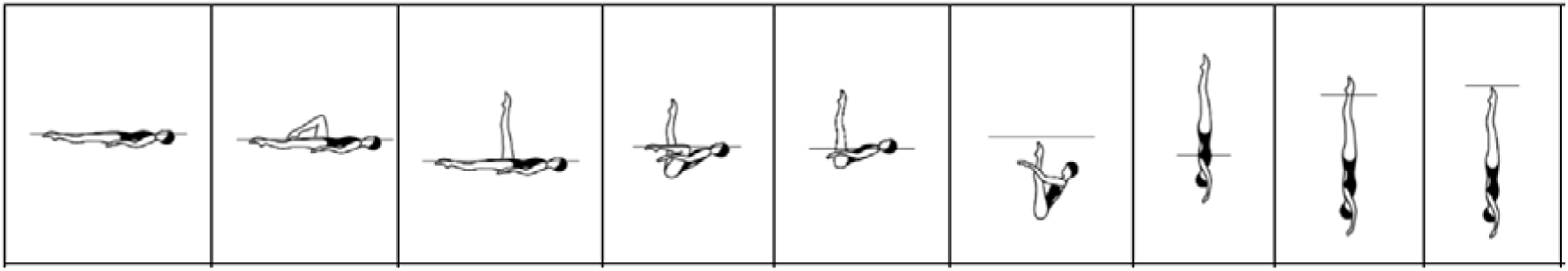
Ariana. World Aquatics. (2022). Figures Manual 2022–2025: Artistic Swimming (Appendix I, p. 2–3). “From a Back Layout Position…Back Pike Position…”

**Figure 2.**
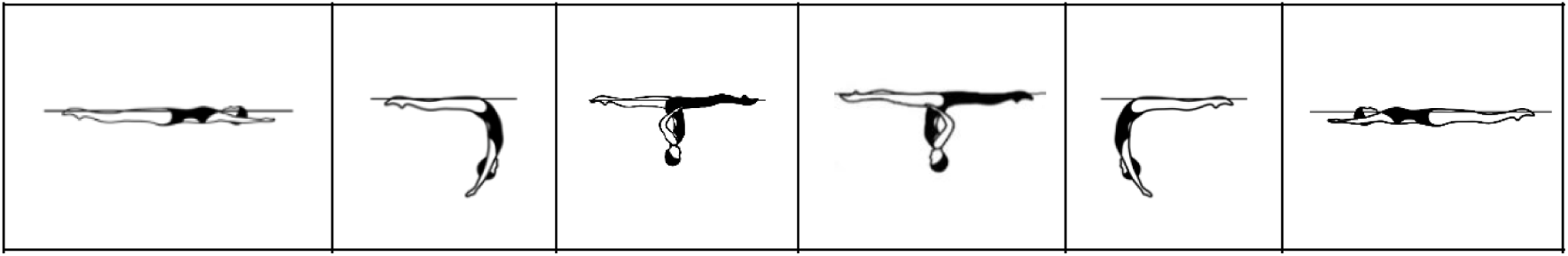
Rio. World Aquatics. (2022). Figures Manual 2022–2025: Artistic Swimming (Appendix I, BM 10 Vertical Descent, pp.lll19,lll33–38).

The athletes were required to be familiar with these figures, as they are mandatory for their category. Three certified AS judges, blinded to the research hypothesis and the group each swimmer belonged to, assessed the figures in accordance with the World Aquatics 2024 rules guide (Table 1) and the average score of the three judges was recorded.

**Table 1.** Obtained points according to the grading for figure execution (World Aquatics, 2024)

| Grading | Points | Grading | Points |
| --- | --- | --- | --- |
| Perfect | 10 | Satisfactory | 5.9–5.0 |
| Near perfect | 9.9–9.5 | Deficient | 4.9–4.0 |
| Excellent | 9.4–9.0 | Weak | 3.9–3.0 |
| Very Good | 8.9–8.0 | Very weak | 2.9–2.0 |
| Good | 7.9–7.0 | Hardly recognizable | 1.9–0.1 |

| **Grading** | **Points** | **Grading** | **Points** |
| --- | --- | --- | --- |
| Competent | 6.9–6.0 | Completely failed | 0 |

### Statistical Analysis

All variables were examined using univariate analysis of covariance, with the baseline measurement acting as the covariate. Effect size (η²_p_) was also computed and presented for all comparisons, and interpreted as small, medium and large for values of 0.01, 0.06 and 0.14, accordingly. The significance level was set at p<0.05and the analysis was carried out using IBM SPSS Statistics for Windows, version 22.0 (IBM Corp., Armonk, N.Y., USA). Data is presented as mean±SD, unless otherwise stated.

## Results

All the descriptive statistics (mean ± SD) regarding the profile of participants (age, height, body mass, athletic experience) for both EXP and CON are presented in Table 2.

**Table 2.** Descriptive statistics (meanL±LSD) for experimental (EXP) and control (CON)

| **Variable** | **EXP** | **CON** |
| --- | --- | --- |
| Age (yrs) | 13.6 ± 0.8 | 13.4 ± 1.2 |
| Height (cm) | 159.8 ± 10.7 | 158.8 ± 9.0 |
| Body mass (kg) | 53.0 ± 3.8 | 51.8 ± 7.3 |
| Athletic Experience (yrs) | 4.4 ± 1.4 | 4.0 ± 1.6 |

There was a significant effect of the Pilates training at POST (when accounting for the baseline scores), with EXP showing improved performance in the split, full bridge and shoulders for flexibility, and improved performance in both sport-specific figures in comparison to CON. No significant difference was revealed for knee flexibility. The mean values, significance and effect size for all variables can be seen in Table 3.

**Table 3.** Flexibility and sport-specific figures scores for both groups (exercising with Pilates (EXP) and the group continuing with usual gym work (CON)), before (PRE) and after (POST) the 8-week intervention. *P* values and partial eta-squared (η²L) for comparisons between groups at POST (when accounting for the baseline scores) are presented. Significance was set at *P* < 0.05.

| Test | EXP PRE | EXP POST | CON PRE | CON POST | P | $\eta^2$ |
| --- | --- | --- | --- | --- | --- | --- |
| Split (cm) | 8.6±6.3 | 5.5±4.6 | 8.3±6.6 | 7.4±5.7 | .028 | .282 |
| Full bridge (cm) | 48.5±17.4 | 37.6±21.6 | 47.0±14.6 | 52.0±15.5 | .003 | .447 |
| Shoulders (cm) | 73.2±20.3 | 64.2±16.3 | 63.1±13.4 | 63.9±15.1 | .005 | .415 |
| Knee flexibility (cm) | 3.8±2.3 | 3.4±2.4 | 2.6±2.2 | 2.8±2.0 | .376 | .053 |
| Ariana (score) | 5.3±0.7 | 5.9±0.8 | 4.6±1.1 | 4.7±1.2 | .001 | .535 |
| Rio (score) | 5.5±0.8 | 6.1±0.7 | 4.6±0.5 | 4.7±0.5 | .003 | .444 |

## Discussion

The results revealed significant improvements in both flexibility and sport-specific figures performance in young female artistic swimmers, following an 8-week Pilates program. Specifically, after the 8-week intervention, EXP demonstrated improved performance in splits, full bridge, shoulder flexibility, and the Ariana and Rio figures compared to CON. The observed improvements in flexibility are consistent with the physical demands of AS activities that require mobility and control (Cho et al., 2017). These findings highlight Pilates’ potential in improving flexibility required for AS, as well as improving the performance of sport-specific figures and consequently improving AS performance.

These findings can have important applications in artistic swimming training regimes. Flexibility is considered the most important characteristic in artistic swimmers (Cho et al., 2017), likely due to its multifaceted impact on performance. Flexibility can have a direct effect on performance, as the figures executed rely on the flexibility of the associated joints to achieve an aesthetically perfect execution (Badau et al., 2021). Consequently, increased flexibility enables a better performance during those figures. Additionally, flexibility has been shown to have a significant effect on motor control in adolescents (Chagas & Barnett, 2023), indirectly impacting and thus cumulatively affecting performance by facilitating better control of execution movement of the figures. This notion is supported by the increased judges’ scores achieved in the sport-specific figures by EXP. Finally, the magnitude of flexibility improvement in present results compare very favorably to the magnitude of improvements seen after a continuous or an intermittent flexibility training program in preadolescent gymnasts (Donti et al., 2018), as well as those achieved in 13-14 year old female artistic swimmers following an ∼8-week flexibility program (Badau et al., 2021). The novelty in the present study is that sport-specific measures were also included to examine whether any increases in flexibility were substantial enough to be translated into performance improvement. Our findings support Pilates as an effective training program, tailored to the needs of AS.

The significance of the present results is highlighted by the fact that in AS, even a minor scoring difference can determine qualification to a final or the awarding of a medal. For example, at the 2024 Paris Olympic Games, the Alexandri sisters of Austria finished fourth in the AS duet competition. They finished with 555.6678 points, missing the bronze medal by only 2.7285 points to the Dutch De Brouwer sisters, who scored 558.3963 points. This difference represents a 0.49% change of the total score, demonstrating how close the competition for the podium was (World Aquatics, 2024). While those athletes compete at world-class competitions, the importance of small scoring differences remains relevant in smaller events. Even in national or regional competitions, minor variations can influence final rankings and qualification results. For example, in a recent Figures Event held in Chania, Greece, the first-place finish was determined by a margin of less than one point, highlighting the importance of these minor changes in the final rankings (Hellenic Swimming Federation, 2024).

Although there is limited literature on Pilates in AS, studies from therapeutic applications and different sports have also reported benefits in performance, after introducing Pilates, as part of their training regimes. In therapeutic contexts, research in children with cerebral palsy indicates that Pilates improves balance, postural control, and trunk function (Abd-Elfattah, Galal, Aly, Aly, & Elnegamy, 2022; Adıguzel & Elbasan, 2022; Panhan et al., 2020), as well as joint stability in hypermobility disorders such as Ehlers-Danlos syndrome (Castori et al., 2017). Pilates also lowers stiffness in hypertonia and improves movement control in hypotonia (Calık et al., 2020; Hornsby & Johnston, 2020). In sporting contexts, Pilates has been found to increase soccer players’ hamstring flexibility (Chinnavan, Gopaladhas, & Kaikondan, 2015, Hägglund et al., 2013). Pilates has also been shown to increase range of motion and muscular strength in dancers, in movements like arabesque and développé, which require core stability and precise control, as well as dynamic posture (Ahearn, 2006; Amorim et al., 2011; McMillan, Proteau, & Lèbe, 1998). It also improves balance, muscle responsiveness, and vertical jump performance in basketball when combined with plyometric exercises (de Costa Ventura, 2020; Panse et al., 2018). Further, it has also been reported to help volleyball players enhance their power and dynamic movements such as leaps (El-Sayed et al., 2010). These studies emphasize the effect of Pilates on the performance of various physical components, particularly those requiring postural control and overall strength.

These findings need to be interpreted with a certain caveat in mind. As indicated earlier, Pilates is a *holistic* training approach encompassing several mental and physical aspects, such as mental and physical control, optimal breathing, flexibility, strength, and precision (Ahearn, 2006; Isacowitz, 2022). While the current study assessed and showed improvement in flexibility, we cannot exclude that the non-assessed, but inherent, other components of Pilates training (e.g. core strength, postural control, motor control, optimized breathing patterns) have also improved. With AS performance relying on good cardiorespiratory fitness, balance, coordination, agility, swimming proficiency, explosiveness and endurance (Chairopoulou, 2010), it is feasible that those components acted in tandem with increased flexibility to contribute to the observed improvements in AS performance. Future studies should aim to elucidate and quantify the individual component improvement and effect on AS performance, so as to provide a more nuanced and complete understanding of Pilates training on AS performance. Regardless of the exact mechanism, the findings support the notion that Pilates training can help improve AS performance.

Despite the positive findings, certain limitations must be acknowledged to fully understand the results and their implications. A drawback of this study is its small sample size, which reduces the likelihood of a statistically significant finding reflecting a true effect and potentially yields erroneously inflated effect sizes (Boyle & Verstegen, 2012). Furthermore, the study used chronological age rather than biological age (Lloyd, Oliver, Faigenbaum, Myer, & Croix, 2014), potentially ignoring developmental or physiological variations that could alter individuals’ responses to Pilates training, as biological age is known to impact performance (Sokołowski et al., 2021; Vandendriessche et al., 2012). Finally, although participants were blinded to the research hypothesis, participants’ awareness of their group assignment, may have in advertently revealed the study’s aim and introduced biases, such as compensating rivalry or resentful demoralization, which could have influenced the results. Despite these limitations, the findings offer useful insights on Pilates’ involvement in AS and lay the groundwork for future research.

The study also offers several notable strengths. The inclusion of a control group, not always possible in ecologically valid training studies (Balasas, Kellis, Christoulas, & Bampouras. 2021), increases our confidence in the observed improvements being directly the outcome of the Pilates intervention. Another significant strength is the utilization of sport-specific tasks, as well as the flexibility measures, that are graded in a way that simulates AS competitive circumstances, enabling translation to real-world performance gains rather than limiting findings to simulated situations or general fitness components. Finally, the implementation of the program for eight weeks provides a realistic time frame both for benefits to be seen as well as for the program to be attractive to coaches.

## Conclusion

This study emphasizes the potential benefits of introducing Pilates into the training program of AS athletes. The results show considerable improvements in flexibility, highlighting the method’s capacity to improve important physical characteristics required for AS performance. Furthermore, these findings resulted in improved execution of figures in water. The absence of significant gains in knee flexibility indicates that the effectiveness of Pilates may vary across different joints and movement patterns. These results support the inclusion of Pilates sessions, at least twice weekly, as an alternative training approach for AS athletes.

Future research should explore two main areas to deepen our understanding of Pilates’ specific impact on AS performance. First, investigating the long-term effects of Pilates on AS, both in the relevant physical components as well as in sport-specific performance, will offer a better insight of the impact and feasibility of implementing Pilates on AS training programs. Secondly, comparing the impact of Pilates with other flexibility-enhancing strategies (e.g., yoga, dynamic stretching) on AS performance could help identify the most effective approaches as well as offer alternative regimes to AS athletes and their coaches.

## Data Availability

All data produced in the present study are available upon reasonable request to the authors

## Acknowledgments

The authors would like to thank the participating athletes and their coaches for their cooperation and dedication throughout the study.

## Statements and Declarations

### Ethical Approval and Consent to Participate

This study was conducted in accordance with the ethical principles outlined in the Declaration of Helsinki. Ethical approval was obtained from the University’s Ethics Committee (Protocol No: 1461/11-01-2023). All participants and their parents/guardians were provided with detailed information regarding the study’s objectives, procedures, and potential risks. Written informed consent was obtained from the parents or legal guardians of all participants, confirming voluntary participation. Participants were assured that their data would remain confidential and that they had the right to withdraw from the study at any time without any consequences.

## Consent for Publication

Not applicable.

## Availability of Data and Materials

The datasets generated and analyzed during the current study are available from the corresponding author upon reasonable request.

## Authors’ Contributions

L.K. and S.N. conceptualized the study and designed the methodology. T.Μ.B. and M.K. assisted in data collection and analysis. E.D. contributed to statistical analysis and manuscript preparation. All authors read and approved the final manuscript.

